# How COVID-19 challenged care for women and their newborns: a qualitative case study of the experience of Belgian midwives during the first wave of the pandemic

**DOI:** 10.1101/2021.05.21.21257440

**Authors:** Elise Huysmans, Constance Audet, Thérèse Delvaux, Anna Galle, Aline Semaan, Anteneh Asefa, Lenka Benova

**Affiliations:** Institute of Tropical Medicine, Antwerp Belgium; International Centre for Reproductive Health, Department of Public Health and Primary Care, Ghent University

## Abstract

In this article, we describe the results of a rapid qualitative study conducted between May 19 and June 25, 2020 on the work experience of midwives during the first wave of the COVID-19 pandemic in Brussels and Wallonia (Belgium). Using semi-structured interviews conducted with fifteen midwives working in hospitals or practicing privately, we investigated the impact of the first COVID-19 wave on their work experience, the woman-midwife relationship, and midwife-perceived changes in quality of care. Findings include high levels of stress and insecurity related to the lack of resources and personal protective equipment, feelings of distrust from midwives towards the Belgian State and public health authorities, as well as structural and organizational challenges within maternity wards which negatively affected quality of care. Moreover, based on the midwives’ experiences, we demonstrate the need to recognise the views of all stakeholders involved in maternal and newborn care provision, and share five essential lessons learned from this study: 1) it is crucial to acknowledge the central role of midwives for maintaining maternal and newborn care amidst the pandemic and beyond; 2) creating unified national guidelines could support ensuring best practice; 3) efforts must be put in place to diminish the climate of mistrust towards health authorities and to repair the relationship between midwives and decision-makers which was damaged during the pandemic; 4) caring for front-line healthcare workers’ mental health is critical, and 5) quality of maternal care can be improved, even in the midst of a pandemic, through team effort and creative solutions tailored to the needs and demands of each setting.

## 1. Introduction

### 1.1. Background of the study

On the 12^th^ of March 2020, Belgium entered a “federal crisis management phase” (1) to counter the spread of the novel coronavirus (SARS-CoV-2). As many other European countries, Belgium has been severely impacted by the COVID-19 pandemic. From the beginning of the ‘first wave’, the Belgian Federal State coordination was conducted by crisis cells composed of different levels of authorities to strengthen communication between the multiple authorities responsible of taking necessary measures to contain the epidemic (2). On the 14^th^ of March 2020, all hospitals entered an “emergency phase” (1). This included increasing the Intensive Care Unit (ICU) capacity and cancelling all non-urgent patient consultations. The resources spared by these measures were allocated to the establishment of COVID-19 treatment units. The changes in the allocation of resources and consultations in hospitals, as well as the need to reduce the spread of the virus, have not been without impact on pregnancy, childbirth and postnatal care. In this article, we describe the results of a rapid qualitative study conducted between May 19 and June 25, 2020 on the work experience of midwives during the first wave of the pandemic in Brussels and Wallonia. Based on semi-structured interviews conducted on online calling devices (Zoom and Skype) with midwives working in hospitals or practicing privately, we investigated the impact of the first COVID-19 wave on their work experience, the woman-midwife relationship, and midwife-perceived changes in quality of care.

Horsche et al. show that few specific data exist regarding the experiences of providers caring for pregnant and birthing women during COVID-19 (3). However, we know that on the international level, the COVID-19 pandemic affected pregnancy care, childbirth and postnatal care from the very onset. Data from as early as March 2020 show that maternal and newborn care workers experienced high levels of stress due to the reduced use of routine maternity care and modifications of care processes, in both high- and in low-income countries (4). Therefore, examining midwife-woman relationships during the periods of pregnancy and childbirth in the era of COVID-19 is imperative — not least because it was challenged by the necessity of using personal protective equipment (PPE) and physical distancing measures. *“Face-to-face psychological support is as important as physical checks, and good quality maternity care requires a trusting relationship between professionals and families. Good eye contact, touch, and tone are critical elements of care, particularly during labour”* (3). Additionally, the philosophy of midwifery and the essence of the specific way of caring requiring frequent physical contact with women is subject to heavy compromise in the COVID-19 context (5). Other data from the United States indicate that maternity care professionals faced a lack of PPE and had unclear guidelines, while partners and doulas were excluded from maternity wards and birthing rooms, leaving women feeling lonely and unsupported (6). Studies conducted during previous outbreaks, such as SARS and Ebola showed the multiple effects of infectious disease epidemics on care provision to patients, and on caregivers’ mental health and wellbeing. Mental exhaustion and anger (7), increasing levels of stress due to the risk of infection (8), and higher risks of burnout and psychosocial distress (9) were among the most frequently reported negative consequences.

Since the beginning of the COVID-19 pandemic in Belgium, several press articles and opinion pieces highlighted negative childbirth experiences of women, perceived as related to the epidemic context (10) (11). They pointed to the difficulties of labouring and giving birth with face masks, the restriction on accompanying persons during childbirth and, more generally, the lack of humanity that resulted from social distancing measures and the wearing of PPE. Additionally, a Royal Decree authorising the Belgian King to take measures to combat the spread of SARS-CoV-2, such as temporarily allowing the practice of nursing by unqualified healthcare professionals (12), gave rise to numerous public debates. One of the targets of these polemics was the recognition of healthcare professions and paramedic professions such as midwifery (13). From a general point of view, Belgium, like many other countries, lacked sufficient quantities of PPE at the beginning of this crisis. In this context, priority in the distribution of masks was given to hospitals and to general practitioners (primary care doctors). Other professionals such as midwives working in hospitals or in the community, had no organized access to PPE and had to cope with the means at hand (2). It is during this “polemical” context of the beginning of the pandemic, including incomplete scientific knowledge on routes of transmission causing frustration and anxiety for health professionals, that we undertook our research with midwives. Through the analysis of one to two in-depth interviews per individual, we summarise the main issues raised about midwifery during the first months of the COVID-19 pandemic.

### 1.2. Maternal and newborn care in the Brussels-Wallonia Federation

Our study focuses on the French speaking part of Belgium (Wallonia and French-speaking part of Brussels), where several healthcare providers are involved in the provision of maternal and newborn care. Women and newborns encounter gynaecologists, paediatricians, and physiotherapists, among others, depending on their needs and insurance coverage, both in health facilities and during home visits. When no pathology that would require a medical intervention is detected, midwives are able to care for both women and newborns throughout pregnancy and thereafter. For “low-risk” pregnancies and childbrith, the length of stay of the mother and newborn in hospitals had recently been systematically reduced in Belgium (from 5 days in 2000 to 3.1 days in 2016 (14)).

In Belgium, midwives can either work in hospitals or as privately practicing providers. Privately registered midwives often provide care at home such as antenatal visits, home births and postnatal care. Midwives who make home visits for these services are recommended either by the hospitals where women plan to give birth, or through personal acquaintances of the families. The pregnant woman is entitled to choose her care professionals during and after pregnancy. In addition, several governmental social support agencies such as the ONE (Office de la Naissance et de l’Enfance) can be involved in supporting women, families and newborns before, during and after childbirth. Services provided by registered health professionals may be free of charge, or reimbursed in whole or in part, depending on the type of service and on women’s insurance coverage.

Midwives carry out pre-conception consultations (medical history, complete anamnesis, genetic counselling, nutritional advice etc.) as well as follow pregnant women throughout their pregnancy. Their role before birth is to conduct basic examinations (blood tests, uterine height, weight control, complete physical examination, gynaecological examinations and ultrasounds in each trimester). Belgian midwives are also competent to prepare their patients for childbirth. Among other aspects, they can provide information, help relieve pain, respond to the concerns of women and their families and prepare for the homecoming with the baby. Additionally, midwives can support births in different health facilities (hospital, birthing centres, etc) or at home, and in different forms such as water birth. In the postpartum period, several visits by midwives in Wallonia and Brussels are usually scheduled within six weeks after birth where a midwife will provide lactation advice, carry out mother and child check-ups, counsel on and provide postpartum family planning, and answer parents’ questions and concerns in various areas. Midwifery is at the crossroads between different health professions, whereby midwives possess a broad and competency-based approach to maternity care (15). Midwives also play an important role as coordinators of care, able to detect anomalies, complications and any event drifting out of the “physiology” of the mother-baby pair and refer them to specialised care if necessary.

However, midwives in Belgium operate within a broader system where the care during pregnancy, childbirth and the postnatal period is considered to be obstetrician-led”, taking place predominantly in hospitals and outpatient clinics (16). In contrast, the midwifery-led care model is applied in the Netherlands, Finland, and England (17). The organisation of healthcare in Belgium is mainly based on specialised medicine and Belgian midwives report being sometimes unable to exercise the full extent of their competencies because of the predominant roles of obstetricians and gynaecologists in pregnancy and childbirth (18). Despite the proven benefits of a midwife-led care model (19), midwives in Belgium are not always considered as the main caregivers for women during pregnancy and childbirth.

## 1. Methods

### 2.1. Study design

This cross-sectional qualitative study used semi-structured online interviews that were audio- and video-recorded. Three core topic areas were explored in the interview guide: practical measures and guidelines introduced in response to the COVID-19 pandemic, the contextual challenges/facilitators of the implementation of the new measures, and experiences of midwives in the first wave of COVID-19.

### 2.2. Study population, settings and samplings

The objective of our study was to report on the main challenges encountered by midwives at the beginning of the pandemic. We conducted in-depth interviews with 15 midwives, interviews ranged between 60 and 90 minutes in duration. Recruitment of respondents relied on a combination of approaches. First, we contacted midwives who responded to the online “COVID-19 Maternity Survey” led by the Institute of Tropical Medicine, Antwerp, were from the Wallonia-Brussels federation and agreed to be contacted by e-mail for in-depth interviews (4). Personal and professional networks of the researchers were also involved to recruit additional respondents. To complete the panel, we recruited respondents via social media (Facebook and Twitter). Finally, one person was recruited during the course of the study through a midwife who had already participated in the interviews.

We interviewed midwives practising privately and in various hospital types and sectors across different regions of Brussels and Wallonia. Three respondents worked in a private hospital, four in a public hospital, five were privately practicing (mainly providing care to women and newborns at home), and three with multiple employment (privately practicing and hospital-affiliated). Altogether, our respondents practicing in hospitals were working in nine different infrastructures. All the respondents were women aged between 20 and 35 years old. None of them was occupying a management position, but one of them occupied a senior role in the Belgian professional organisation for midwives (UPSFB).

Given the social distancing and lockdown measures in place during the study, two of the researchers (EH and CA) conducted interviews via online platforms (mainly Zoom and Skype). We exchanged e-mails with the respondents in order to agree on a one-and-a-half-hour slot for the semi-structured interview. We sent them the main topics of the discussion beforehand but did not give them our interview guide so that we could adapt our questions to their answers. The researchers were at their respective homes. Depending on their family context, the midwives were at home alone or with family in the same room (but not participating in the interview). More broadly, we tried to find quiet moments when our interlocutors could express themselves as freely as possible about their midwifery activity.

At the beginning of each interview, the researchers collecting data introduced themselves, explained the objectives of the study and the ethical obligations concerning the study, especially to warrant anonymity and confidentiality and asked for verbal informed consent. Interviews were recorded with the permission of each respondent, and these recordings were deleted after transcription, ensuring anonymity and confidentiality and establishing a framework of trust for our respondents.

### 2.2. Data collection

Each respondent was interviewed once or twice, depending on the context. One midwife asked for more time to fully go over her complex situation and we agreed with her to make another appointment to cover more topics that we wanted to discuss together. After a follow-up interview with another midwife during the month of June 2020 and after contacting several other respondents to make a second appointment, we realized they did not report any updates about their work situation at that time. In addition, with the decision to loosen the lockdown measures around May and June 2020 and the end of the state of emergency in hospitals, our interlocutors expressed to us a feeling of a “return to normal”. Therefore, we only interviewed two respondents twice. The rest of the respondents (n=13) were interviewed once. We decided, given the objective of covering the time of the first peak of the pandemic, that we had reached saturation of data for our study in June 2020.

The interviews were conducted in French (the mother tongue of the researchers and all the midwives) and then translated into English by the researchers, supported by English speakers from the research team at the Institute of Tropical Medicine, Antwerp. The two researchers in charge of the interviews took turns leading the respondent interviews, the other interviewer taking on the role of “observer” and ensuring that the technical details went smoothly (registration, proper functioning of the online platform, etc.). In addition, as the interviewer duo consisted of a medical doctor (CA) and a health anthropologist (EH), they each focused on questions in their specific field of research.

We used a semi-structured interview guide. This was then adapted into a practical visual highlighting the main themes to be addressed. The questions were then adapted during each interview according to our interlocutors. The purpose was to be able to exchange with our respondents on some concerns we originally did not address in our interview guide. If necessary, the guides were adapted between interviews when the researchers felt that a question was not understood, or when certain themes were found to be missing during the early stages of data collection.

### 2.3. Analysis and inductive method

Data were analysed using an inductive thematic analysis method. Throughout the data collection and analysis, we proceeded by iteration, going back and forth between data collection, analyses and revisiting our research questions, each of these three components having a non-linear influence on the others (20). We cross-referenced the information collected in order to take into account the heterogeneous and contrasting discourse of the interviewees, notably by using a spreadsheet for cross-analyses between the narratives of each person and the identified themes. Coding and reviewing were done by both interviewers (EH and CA).

### 2.4. Reflexivity and positioning of the researchers

This study benefited from a multidisciplinary collaboration between medical science and anthropology in both data collection and analysis. The analysis was also done in collaboration with a broader team including a Belgian midwife, a Belgian gynaecologist/obstetrician, a nurse and two maternal health researchers. Given that the problem is both medical and social, we felt it was impossible to ignore a process based on a combination of methodologies and points of view. As Fainzang clearly puts it: “*One of the contributions of anthropology lies in its ability to rethink and redefine the categories used by medicine, and to free these categories from the content assigned to them by medicine, in order to question their social significance (…) Because research issues (like health issues) are never exclusively health issues, but also always social, political and economic issues at the same time*.” (21). Throughout this research, we have endeavoured to maintain this plural and global vision of what midwives’ experiences during the COVID-19 pandemic are based on. As Hermesse et al. (22) explains, social sciences make it possible to analyse the diversity of responses to the pandemic according to the lives and contexts of each person. Moreover, anthropology is particularly concerned with understanding how this coronavirus modifies our togetherness and social interactions (22). How have we looked at and considered others since the beginning of the pandemic?

### 2.5. Ethical considerations

This study was approved by the Institutional Review Board at the Institute of Tropical Medicine, Antwerp on May 5, 2020 (reference 1372/20). Throughout the manuscript, the names of the midwives were changed in order to protect their anonymity.

## 2. Results and interpretation

We identified four key themes during this rapid qualitative study. First, we present how the new guidelines in times of COVID-19 were perceived by the midwives and affected their day-to-day professional lives. Second, we describe how the disruption in the organisation of care was identified as a major issue for midwives. Third, we introduce how the midwives reflected on their work experiences during the first wave of the COVID-19 outbreak in Belgium. Finally, we describe, based on the discourses of our informants, the perceived changes in the woman-midwife relationship.

### 3.1. Establishing guidelines for provision of maternity care in times of COVID: the age of resourcefulness?

In the fight against SARS-CoV-2 transmission in Belgium between March and June 2020, hospitals established their own guidelines independently. These guidelines included both routine care and care for COVID-19 positive pregnant women. At the time of the study, little was known about the effect of COVID-19 on pregnant women and experiences and views of maternal and newborn healthcare workers had not been empirically documented yet (4). This led to major discrepancies in rules and guidelines between hospitals. Furthermore, midwives reported not always being aware of team/individuals in their hospitals responsible for establishing the measures against coronavirus disease transmission, leading to confusion among healthcare providers in the wards. Noémie, a midwife working in a public hospital, describes: “*The instructions we had in the maternity ward given by the gynaecologists were not the same as those we had been given in the delivery room (*…*) and the paediatricians in neonatology did not say the same thing as the paediatricians in the maternity ward. It is not clear who has to make the decision (*…*) and I don’t know who to talk to anymore* [about the new guidelines]”.

When a group of experts was designated to manage the COVID-19 response in a hospital, some respondents mentioned disagreements within the medical team, leading to a sense of lack of unity and coherence. Among the nine hospitals where our respondents were working, none was described as including the midwives in the decision-making process and establishment of guidelines for maternity care. As Béatrice, a midwife working in a public hospital, illustrates: “*There is a reflection at their* [the gynaecologists’] *level, between them, we know they talk a lot in their WhatsApp group, but we don’t know anything about what comes out of that. They sometimes make decisions, but they never consult us (*…*) or ask for our opinion”*. The feeling of marginalisation for the midwives within this process regularly led to frustration of not being recognised as a central actor in provision of healthcare to women and newborns and led to decisions that were not aligned with midwives’ work values.

In most hospitals, and in the early stage of the pandemic, guidelines were changing several times a day, creating various challenges. Midwives lacked time to get acquainted with the updates and many reported a feeling of discouragement by the frequency of the updates, despite understanding the need for regular updates as new evidence were constantly emerging. The quantity of information to be remembered dramatically increased midwives’ workload as well as their mental load. The lack of clear and summarised guidelines also impacted the midwives’ feeling of safety. In fact, midwives expressed the fear of not being sufficiently prepared either to provide safe and high-quality care to their patients, or to protect themselves from infection. Caroline, a midwife in a public hospital, describes: “*The folder with the procedures changed constantly in the first month, we always had to check for the updates on how to care for COVID positive patients and it took us a lot of time to feel safe when caring for COVID patients, as the procedures were changing all the time”*.

Guidelines for preventing the spread of the coronavirus for privately practicing midwives were issued by the professional union of midwifery (UPSFB) at the end of March 2020. The information and guidance concerning home-visits were made available by the professional union of midwives on their closed Facebook group but made sure to display them on their website as well to reach a broader audience, making information accessible to midwives outside of their union. Many challenges had to be overcome by midwives working outside of hospitals. Some respondents highlighted how they had to be creative in adapting the guidelines to their daily work. As Rebecca, a privately practicing midwife describes: “*The word for that period is really “resourcefulness”! And every day I went to see people, every day I had a new thing to manage and I thought “How am I going to do it?”, I found solutions but well*… *I really had to be creative!*”. Several examples demonstrate the creativity of midwives and their daily “tinkering” in the face of a situation that was perceived to be unstable and not meeting their needs. Some midwives sewed their own face masks or had them sewn by family members or friends. Others tried to recover stocks of PPEs from other industries, such as industrial construction, which had a surplus of FFP2 at that time. Others living near the French border bought face masks from pharmacies in France to redistribute to their colleagues. Some even went as far as to check the lists of medical professionals entitled to the distribution of PPE. By reporting the names of deceased health professionals, they requested to obtain the masks intended for them. For home visits, some dressed in garbage bags to cope with the lack of medical aprons.

#### 3.1.1. Access to PPE at the core of the feeling of insecurity and distrust of the public authorities

As we can see, for most healthcare workers during the first wave of the pandemic, midwives’ access to PPE, especially surgical face masks, was one of the biggest challenges in the early management of the COVID-19 crisis (23) (24) (25). We could identify three pillars to the feeling of insecurity that arose with the restricted access to PPE for midwives: fear of transmitting coronavirus to patients, fear of getting infected themselves, and fear of bringing the virus home and putting their families at risk.

The distribution and recommendations regarding the use of face masks for healthcare workers outside of the COVID-19 wards at the time of the study were unclear and lacked organisation (2). As we already noted, some midwives reported defying these recommendations by sewing cloth masks or wearing and washing surgical face masks at home in order to feel safer, as Adélaïde explains: “*Every evening I came home, washed my mask and went back to work the next day with it. There we went from insecurity to anger because I was furious! (…) I thought it was absurd, I felt like I was doing humanitarian work when I’m in one of the biggest university hospitals!*”

Adding to the feeling of vulnerability against the new virus, a strong feeling of distrust and anger arose among midwives toward their hospitals and the government for the mismanagement of PPE stocks. Several of our respondents were told that guidelines recommended the use of masks at all times but not enough of them were available. This situation played an important role in the mental load and stress experienced by midwives at this time.

The same issue was also reported by privately practicing midwives outside hospitals. At the very start of the pandemic, there were no PPE accessible for healthcare workers outside COVID-19 wards (2). Local municipalities in Belgium still tried to distribute face masks to other essential and privately practicing healthcare workers but our informants working in private practices or doing only home-visits reported feeling excluded from this local distribution process. Marie, from the professional union of midwives, took part in the distribution of masks and reported several frustrations: “*The lists* [of health providers] *were not correct. We sent the complete listing to the health authorities a dozen of times (*…*). Then, there was also the transportation of the masks from the authorities to the municipalities. In the end, the army had to deliver our masks! That’s crazy! They lost entire trucks*!”. This created a lot of frustration and feelings of distrust among privately practicing respondents. As Nathalie reports: ‘*It’s hard to say as a professional: “I can’t care for families because I’m not given the equipment, I need to protect myself and my family*”. Additionally, the absence of midwives on the priority lists raised questions about the response capacities of the Belgian health authorities in times of crisis, as well as the status and recognition of midwives in the medical establishment altogether.

#### 3.1.2. Testing strategy: improving security or reinforcing the feeling of mismanagement?

Every hospital was responsible for organising its own SARS-CoV-2 testing strategy for pregnant women coming for childbirth. These testing strategies in several hospitals were reported by the midwives to drift away from the national recommendations in terms of target population and methods, although the Polymerase Chain Reaction test (PCR) test of a nasopharyngeal swab seemed to be the most common test used. Some strategies were often seen by midwives as profit-driven for the hospitals, based on fear, or simply inefficient. Some hospitals were asking women to only get tested 2 weeks before the due date, a strategy that raised questions for our informants, as women return home and may become infected in the period between the test and delivery, making the test result unreliable and putting healthcare staff at risk on admission. The interviewees raised concerns about some recommendations that could lead to malpractice and some measures taken could be perceived as violence against women. As described by an independent midwife: “*I saw absolute nonsense! (*…*) In fact, it was financial. Every pregnant mother had a CT-scan to make sure she was not COVID positive. Or hospitals where (*…*) mothers were tested at 38 weeks, if the test was negative, women were induced within 48 hours to be sure* [they would not get infected]. *I hear all the fears and anxieties, but for me it still sounds like violence*”. Additionally, many midwives deemed that health facilities were not doing everything possible to protect their employees, contributing to further exacerbate the feeling of distrust towards authorities in charge. Adélaïde reports: *‘We’re considered as cannon fodder, when it* [the patient’s test] *is positive we’re not even called to get tested, clearly there’s something wrong about the procedure*”.

### 3.2. From structural to organisational challenges: how COVID-19 affected maternity wards

The general organisation of maternity wards evolved as the pandemic expanded. Once again, a unified guidance for hospitals in the Brussels-Wallonia Federation was absent, and we could document many different ways to organise care. The main recommendation for all healthcare professionals during the pandemic was to cancel all non-urgent outpatient consultations and reduce contact. Earlier discharge after childbirth was also common, so that less care would be provided inside the hospital. The midwives working in hospitals reported that the decision about the discharge time was made by the clinical team (midwives, obstetricians and gynaecologists, and paediatricians) in agreement with each individual woman and based on her readiness. However, some privately practicing midwives reported situations where new mothers were discharged from a hospital as soon as 24 hours after birth, without being consulted. As Nathalie mentions, independent midwives were actually not always feeling prepared to care for women at home so soon after birth, especially with the difficulties to procure PPE at the beginning of the pandemic: “*Getting people out 24 hours after giving birth is quite a big deal, especially in a pandemic like this, and not finding the equipment, I found it quite anxiety provoking”*.

In most hospitals, the pandemic affected the clinical decisions taken by medical teams, with the aim of shortening hospital stays. The midwives reported that the usual obstetric guidelines were disrupted, which could create conflicts between the medical teams and both patients and midwives. Nathalie, working part-time in a public hospital and part-time privately describes: “*When their water breaks (*…*) normally we don’t induce the birth and we let labour begin. But with COVID, as soon as they arrived in the hospital with a ruptured membrane, they* [the gynaecologists] *wanted to induce childbirth. The mothers didn’t agree, it created conflicts with the gynaecologists, but (*…*) they* [the mothers] *felt heard, and the gynaecologists agreed”*.

In a couple of hospitals, skin-to-skin contact with the newborn was not encouraged when mothers had positive COVID-19 test results. This seemed inconceivable to most midwives. As Kim, working in a private hospital, expresses: “*For example, skin-to-skin was controversial and today it is still not clear (*…*) it was terrible for these patients. We didn’t know how to explain it to them, it was very complicated. Even for us, because saying to a lady, “Listen, you’re going to live with your baby, but you can’t do skin-to-skin”. It was difficult for them and for us, and really against our value*s”.

### 3.3. Midwives’ work experience during the first peak of the pandemic: reconciling health measures and work values

The clinical guidelines in place at the time of the study did not fully cover the complexity of the situation for maternal and newborn care at home and in hospitals, and often overlooked the psychological, social and economic aspects of midwives’ work and relationships with their patients. Hence, many midwives expressed the challenges they encountered when trying to apply some of the measures in the maternity wards or when taking care of women in their homes. Some measures, such as banning birth companions or instructing women to labour and deliver while wearing a surgical face mask, contradicted midwives’ “values”. One midwife testifies: “*if you had the misfortune of being COVID-positive, birth companions were not allowed in the delivery room you had to give birth alone. That’s just awful! We lost our humanity!*”

As a result, most of the midwives reported bending the rules to ensure the implementation of what they considered as non-negotiable elements of respectful care to women and their babies. Adélaïde described the situation in the maternal intensive care unit: “*We made a lot of exceptions to preserve the family unit. We lied about the reasons for staying in a single room, saying it was for medical reasons when it wasn’t*… *because we needed it, it was our way of compensating for separating other couples*”. She continues: “*We’ve made exceptions sometimes. For the ladies who had been here for over two months, we allowed the husbands to come for one afternoon. The head midwife agreed with that, but we organised the visit in the patients’ kitchen, we closed the door so that the other patients wouldn’t see, so it was a bit clandestine!* (…) *At some point, we all broke the hygiene measures a bit for the psychological well-being of the ladies*”.

To witness the drastic measures taken to prevent the virus transmission without being consulted had a major impact on midwives’ levels of stress and mental load. However, as the pandemic situation evolved, the midwives reported that hospital hierarchy became more receptive to their viewpoints and demands and agreed to adapt some measures. By giving feedback, midwives’ experiences improved. They started to feel heard, their views were taken into consideration and were better able to care for women according to the principles of their profession.

The infection prevention and control measures had several consequences on healthcare providers’ everyday activities. In hospitals, donning and doffing of PPE, and planning care procedures differently to avoid back-and-forth inside isolation rooms for COVID-positive women significantly added to their mental load. The respondents also observed that not entering isolation rooms frequently prevented them from providing the same level and quality of care as they provide for COVID-19 negative patients and isolating the families even more. Sophie explained the difficulty she faced when patients were in isolation: “*I sometimes had the feeling of abandoning my patients* [in isolation rooms] *because those were patients that we went to see less spontaneously*”. However, we should note that few maternity hospitals had, according to the midwives, been taking care of more than four COVID-19 positive women at any one time. Some midwives from our study even never encountered a situation where they had to care for a woman infected with the SARS-CoV-2 virus.

### 3.4. Maintaining a relationship in times of social distancing and PPE: how COVID-19 impacted the patient-caregiver bond

The disruption caused by the pandemic was visible in the organisation of care itself but was particularly evident in the individual relationship between women and their medical providers. As mentioned before, the connection between the woman and the midwife is unique as it entails the weaving of a trusting relationship throughout the woman’s pregnancy, in a continuity of care model. Therefore, and particularly concerning midwifery, many obstacles were faced with the measures taken against COVID-19, including social distancing and the use of PPE.

Midwives reported having more difficulties creating a strong bond with women when wearing full PPE because of the physical barrier this imposed, making care more impersonal as patients were not able to see the faces of their providers. Moreover, midwives felt less able to detect the emotional distress of their women due to the added mental and workload as well as the shortened time spent with women. Sophie, working as an independent midwife and in a public hospital, describes her experience with PPE: *“The few people* [the mothers] *are seeing* [while at the maternity] *are invisible, unrecognisable (*…*) it was completely anonymous care and it was complicated in these moments* … *so intense at the beginning of a child’s life (*…*) to have all these physical and emotional barriers (*…*) because there are so many things that go through the eyes and the non-verbal, it was extremely complicated to sometimes establish a link with our patients. That’s a really difficult thing to do. Especially for a job where we are so much in touch with the human being, in the exchange (*…*) Even gloves, touch is so important, the touch of the babies too! It’s a very, very big barrier, much more important than one could imagine*.*”*

Furthermore, visits of mental health professionals in the maternity wards decreased drastically, together with those of a companion, exacerbating the feeling of loneliness among patients, as Lou describes for the maternal intensive care: “*Usually there is always a friend who comes to spend the day when the husband is working*… *Here we have high-risk pregnancies, medical abortions, miscarriages… all alone! (*…*) We are present, we are there, but the caregiver is not enough in the whole grieving or healing process (*…*) and we usually have the psychologist visiting, and here it was only for selected situations, and I don’t know if we took the time to pay attention to all that…*”. Not always being able to attend to the women’s psychological distress added an important emotional stress among midwives. The loneliness of women in maternity wards, both physically and psychological, the despair of giving birth in times of crisis, and the stress of the crisis itself were considered extremely difficult to face for midwives working in hospitals.

Conflicts between health professionals and patients arose due to infection prevention and control measures. Reasons were diverse, from the anxiety of being in a hospital to the inability to go out of the room to smoke. Béatrice, a midwife working in a public hospital, reports the situation she encountered: “*There’s a husband who panicked because he saw a cart with all the isolation equipment* [PPE] *in front of a room, so it scared him very, very much to think that we were taking care both of his wife and of someone who might be positive*.” Midwives were made responsible for enforcing these rules about visitors, assuming a policing role within their ward; a part which midwives were sometimes not comfortable playing: “*We are not supposed to be in prison in a hospital*”, continues Béatrice.

The loneliness of women was also witnessed by midwives doing home visits, which led them to accept that some of the national social distancing recommendations were not adhered to by families: *“I must admit that several times, with several mothers, I saw that just before I arrived, they made the grandmother come out quickly! I didn’t say anything, I told myself that it was human! (*…*) I saw one of them running away once, I said to myself “she must have seen me arrive, she left quickly”“*, says Rebecca, a private practitioner.

Midwives visiting at home were sometimes the only healthcare professionals that those families were seeing in the postnatal period. Chloé, an independent midwife, faced this situation on several occasions: “*I was stressed because I felt like I was going more* [to patients’ home] *than usual. Talking with different colleagues, everyone had the same conclusion, in fact we have no choice, because they only had us (*…*) I was their only social link*, [they were] *rather very welcoming, but I had to limit the visits after a while, I know it’s nice for them, but I have to be able to justify them too*^*1*^”.

## 3. Limitations of the study

As for all forms of research, but especially in inductive research investigating the views of “others” (26), we acknowledge that our study relies, among others, on the nature of the connections between the researchers and the informants. Given the context of unprecedented crisis at the time of the study, we noted a need among our informants to make sense of the new events that were transforming their professional lives. They reported having little space to express their frustrations, and that the interviews with the researchers allowed them to share their dissatisfactions at work. The particular connection between researchers and informants that resulted from this context was taken into account in our analyses. The fact that our informants focused on the negative aspects of their work is a bias to acknowledge but at the same time, this type of connection was essential to reflect on the few opportunities our informants had to express dissatisfaction in their professional lives.

All interviews were conducted via online videoconferencing. While this was necessary given the lockdown measures in place and can have many practical advantages in terms of saving time and money (27), we recognize that this study is limited to the discourse of our informants and not their practices. To limit this bias, we asked the midwives to keep a logbook and offered to contact us via text or voice messages on the WhatsApp platform. However, this offer was not taken up by the midwives due to lack of time on their part.

A more in-depth study, with the possibility of observing the daily practices of midwives (e.g., an ethnographic study) and an expansion of the circle of informants would make it possible to nuance and clarify the issues in progress for midwives during the COVID-19 pandemic. This study, although limited in time and data collection, does however demonstrate the need for more in-depth studies on the subject.

## 4. Conclusion

The disruptions caused by the first wave of the COVID-19 pandemic for maternal and newborn health in Brussels and Wallonia were numerous and not all yet known nor their extent. In times of pandemic, the need for rapid responses to ensure the continuity of maternal and newborn care justifies the search for concrete and practical solutions to avoid the spread of the virus through routine care performed by midwives. “Learning hour by hour” is a reality rarely experienced by Belgian midwives before the COVID-19 pandemic and strong leadership is needed to reassure the maternity teams and reduce occupational stress, as it has already been shown in the UK (28). Our rapid study allowed us to gain a deeper understanding of the experience of midwives during the first wave of COVID-19 in Belgium and to put this into the context of a profession which remains poorly recognised on a national level. The respondents also expressed the need for better consideration of support and management of mental health for care providers. Poor mental health and increase of occupational stress tend to greatly affect the quality of care and overall wellbeing of the patients and caregivers (29). This issue is seen as a long-term consequence of the COVID-19 crisis for midwives and other healthcare professions, making it a significant part of the concerns of the “aftermath” of the pandemic (5).

As a result of our study and Belgium’s particular context in terms of maternal care, we were able to observe the challenges of the mothers’ and newborns’ transition from hospital to home. The lack of communication between privately practicing midwifes and hospitals made the transition more complex. In both settings, in hospital and at-home, it seemed more important to have a strong communication system between home providers and hospital providers and reinforce the continuity in the care to ensure a good quality of care, through clear and adapted guidelines. Our findings, along with similar researchers in Europe, stress the need to develop high-quality care based on grounded-evidence and co-created with the various actors involved in maternal and health (29). Therefore, we summarized five essential lessons based on Belgian French-speaking midwives’ experiences:

1. <u>Acknowledging the central role of midwives, amidst a pandemic and beyond</u>: concerning maternal and newborn health, our study shows a need to improve midwives’ recognition in Belgium. As seen during the COVID-19 crisis, the discrepancies between decision makers and care providers as well as the lack of recognition of midwives as essential first-line workers, created a gap in the continuity of care, which could be harmful for the women and their babies.
2. <u>Ensure best practices</u>: our findings around the lack of coherence in maternal care guidelines and measures applied by hospitals calls to question how unified national guidelines could be followed by hospitals to the extent of their abilities. Especially in times of disruptions to healthcare provision, the emphasis should be put on ensuring and maintaining the quality of care, defined by WHO (23) as “safe, effective, timely, efficient, equitable and people-centred”.
3. <u>Rehabilitating trust towards authorities</u>: midwives’ feeling of disempowerment created a climate of distrust. There is a demand that policymakers, at national or hospital level, be more transparent about risk management, in order to guarantee a feeling of safety for both caregivers and patients. Communication is seen as a pillar in the management of a health crisis at all levels and midwives report a need to avoid misinformation to ensure continuity of care.
4. <u>Caring for mental health</u>: For both healthcare workers and families, mental health is an integral part of care pathways. Our findings show that the health crisis has led to a major psychological pressure for midwives, compromising their ability to provide quality care at the level that they are accustomed to and that matches their values.
5. <u>Need for creative solutions</u>: Working on the clinical front-line, healthcare professionals have a unique insight of the challenges encountered when trying to maintain quality and respectful care. This calls for tailored solutions adapted to the needs and demands of the population.

## Data Availability

Data cannot be shared publicly because they could compromise the professional lives
of the participants and cannot be fully anonymised. Data are available from the Institute
of Tropical Medicine, Antwerp Ethics Committee (contact via) for
researchers who meet the criteria for access to confidential data.

## 6. Acknowledgements

We would especially like to thank the midwives who answered our questions, who took the time to remember and share difficult moments in their professional and personal lives.

## Footnotes

1 During the first wave of the pandemic in Belgium, midwives could only personally see patients at home when absolutely necessary and were asked to justify their visits to health authorities.

## Notes

### Competing Interest Statement

The authors have declared no competing interest.

### Funding Statement

This study was funded by the Institute of Tropical Medicine's COVID19 Pump Priming
fund supported by the Flemish Government, Department of Economy, Science &
Innovation.
https://www.ewi-vlaanderen.be/en
Grant number: 756141
Dr Benova is funded in part by the Research Foundation, Flanders (FWO) as part of
her Senior Postdoctoral Fellowship.
https://www.fwo.be/
Grant number: FWO / 196000 / LG
The funders had no role in the study design, data collection and analysis, decision to
publish or preparation of the manuscript.

